# Assessing the feasibility and effectiveness of household-pooled universal testing to control COVID-19 epidemics

**DOI:** 10.1101/2020.10.03.20205765

**Authors:** Pieter Libin, Lander Willem, Timothy Verstraeten, Andrea Torneri, Joris Vanderlocht, Niel Hens

## Abstract

Current outbreaks of SARS-CoV-2 are threatening the health care systems of several countries around the world. The control of SARS-CoV-2 epidemics currently relies on non-pharmaceutical interventions, such as social distancing, teleworking, mouth masks and contact tracing. However, as pre-symptomatic transmission remains an important driver of the epidemic, contact tracing efforts struggle to fully control SARS-CoV-2 epidemics. Therefore, in this work, we investigate to what extent the use of universal testing, i.e., an approach in which we screen the entire population, can be utilized to mitigate this epidemic. To this end, we rely on PCR test pooling of individuals that belong to the same households, to allow for a universal testing procedure that is feasible with the current testing capacity. We evaluate two isolation strategies: on the one hand *pool isolation*, where we isolate all individuals that belong to a positive PCR test pool, and on the other hand *individual isolation*, where we determine which of the individuals that belong to the positive PCR pool are positive, through an additional testing step. We evaluate this universal testing approach in the STRIDE individual-based epidemiological model in the context of the Belgian COVID-19 epidemic. As the organisation of universal testing will be challenging, we discuss the different aspects related to sample extraction and PCR testing, to demonstrate the feasibility of universal testing when a decentralized testing approach is used. We show through simulation, that weekly universal testing is able to control the epidemic, even when many of the contact reductions are relieved. Finally, our model shows that the use of universal testing in combination with stringent contact reductions could be considered as a strategy to eradicate the virus.

## 1 Introduction

The SARS-CoV-2 pandemic has caused over 10 million COVID-19 cases and over 0.5 million deaths around the world, since September 2020 [1]. This infection count is presumably an underestimate due to the large proportion of asymptomatic cases [2]. While there are presently different vaccine and treatment candidates being evaluated in clinical trials [3, 4], the control of SARS-CoV-2 outbreaks currently relies on non-pharmaceutical interventions. Whereas, at the start of the pandemic invasive measures such as a full societal lock-down were used to avoid an overflow of the intensive care units [5], currently, many countries aim to control their local SARS-CoV-2 epidemic using a combination of social distancing, teleworking, mouth masks and contact tracing. Yet, while these measures have the potential to reduce the number of detectable infections below 20 cases per 100.000 individuals per day^1^, this still leaves regions prone to local outbreaks, that again require more stringent mitigation measures with societal and economical implications. Since September 2020, many countries had problems controlling the virus, including Galicia (Spain) [6], Antwerp (Belgium) [7], Israel, and the United Kingdom.

The burden of hospitalisation and COVID-19 related mortality seems to be the major motivation to reduce the number of infections. However, keeping the number of infections as low as possible is in the overall population’s interest, considering recent reports on COVID-19 related morbidities throughout all age groups, including neurological conditions, persistent post-recovery symptoms, cardiac injury and pulmonary fibrosis [8, 9, 10, 11]. However, as pre-symptomatic transmission remains an important driver of the epidemic, it comes as no surprise that contact tracing struggles to fully control SARS-CoV-2 epidemics [12, 13, 14]. This is further complicated by the fact that contact tracing is sensitive to the reported number of contacts, which depends on the reporting compliance of the traced individual [13, 14]. Recently, the use of universal testing (i.e., testing the entire population of a geographical region) has been suggested as a solution to suppress SARS-CoV-2 epidemics [15, 16]. Yet, the number of tests necessary to test a country’s entire population in a reasonable time window, remains a serious impediment to this approach.

In this work, we use PCR test pooling, i.e., we combine a number of samples into a pool and test this pool using a single PCR test. As it is logistically most convenient to test household members at the same time, we construct pools out of individuals that belong to the same household. Additionally, testing household members simultaneously agrees with the fact that household members are prone to infect eachother [17]. We consider sample pooling for pools of size 16 and 32, for which recently PCR test sensitivity scores were established [18].

This approach facilitates two isolation strategies. Firstly, *pool isolation*, where we isolate all individuals that belong to a positive PCR test pool, regardless of their individual infection status. Secondly, *individual isolation*, where we determine which of the individuals that belong to the positive PCR pool are positive, through an additional testing step. In this isolation strategy, the individuals in the positive PCR pool are kept in isolation until the individual testing results become available, upon which the individuals that test negative are released from isolation. Thus, in the individual isolation strategy, only the individuals that are responsible for the positivity of the pooled sample are isolated. Both isolation strategies have their advantages and disadvantages. On the one hand, when each individual that belongs to a positive pool is isolated (i.e., pool isolation), this means that negative individuals will be isolated as well, which might have implications for the community compliance with respect to isolation. The pool isolation strategy reduces the need for additional tests, rendering it a preferred strategy when the prevalence is high, and the required number of additional tests is unavailable. On the other hand, when prevalence is low, the number of additional required tests is expected to be low as well, and in such an epidemic phase, performing the additional tests necessary for the individual isolation strategy might prove worthwhile, as it could increase isolation compliance.

For both strategies, it is necessary that the pools have a similar number of households, i.e., that the difference in number of households between pools is minimal, to minimize the number of households that are affected when a pool tests positive. To meet this objective, we devise a heuristic allocation algorithm to assign households to a set of pools of equal size.

We evaluate this universal testing approach in an individual-based epidemiological model in the context of the Belgian COVID-19 epidemic [19] and demonstrate that it is possible to test the whole Belgian population (11 million individuals), in a time span of 1 to 4 weeks. For this, we rely on the projected testing capacity that will be available in Belgium by autumn [20], to accommodate the expected increase in respiratory infections [21]. While we conduct experiments concerning the implementation of universal testing to test the whole Belgian population, we note that the presented framework can also be used to design reactive policies to control local outbreaks (e.g, cities) [22]. In order to assess the robustness of our universal testing approach, we consider different levels of testing and isolation compliance. Furthermore, we consider different false negative rates of the pooled PCR test and the impact of the pool size.

In this work, we show through simulation, that universal testing is able to control the epidemic, even when many of the contact reductions are relieved. Additionally, universal testing implicitly implements surveillance at a high resolution, resulting in a good estimate of the actual incidence and the heterogeneity of this incidence with respect to geography and age. This detailed view on the state of the epidemic will ensure that emergency signals are picked up more rapidly, enabling a swift response that might avoid more invasive control measures.

We acknowledge that the implementation of universal testing is challenging to organise. Therefore we discuss the different aspects related to sampling and PCR testing, to demonstrate the feasibility of universal testing when a decentralized testing approach is used. Finally, our models show that, in the event that a vaccine would not become available in due time, the use of universal testing in combination with stringent contact reductions, could be considered as a strategy to eradicate the virus.

## 2 Methods

### 2.1 Household-to-pool allocation

Consider a set of households ℋ, where each household *h ∈ ℋ* is a set that is comprised of individuals, i.e., household members. From ℋ, we have the total population size,

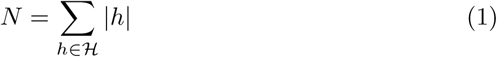

We aim to allocate the households to a sequence of pools with a fixed capacity *k*:

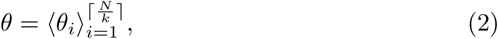

where each pool *θ*_*i*_ is a subset of ℋ, for which

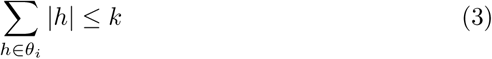

As stated earlier, we aim to construct pools with a similar number of households, and thus our objective is to minimize the difference in number of households between pools:

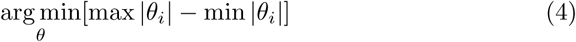

To this end, we formulate a greedy^2^ heuristic algorithm (Algorithm 1). First, the households are sorted in a descending order according to their size, resulting in a set ℋ_*s*_. To initialize, we construct an allocation *θ* of empty pools. Then, in each step of the algorithm, we take away the top item of ℋ_*s*_ and add it to one of the pools that has the least households and members. Note that, to ensure a realistic logistic setting, this algorithm should be run on a set of households that live in close proximity. As detailed in Section 2.3, we run this algorithm on households that belong to the same province (i.e., an administrative region in Belgium).

From the most fundamental perspective, this algorithm always adds a household to one of the pools that has the least number of households, which is a heuristic that will result in an optimal allocation with respect to the objective stipulated in Equation 4. This heuristic enables us to create an optimal allocation for the populations considered in our experiments (Section 3).

#### Algorithm 1: Household to pool allocation algorithm

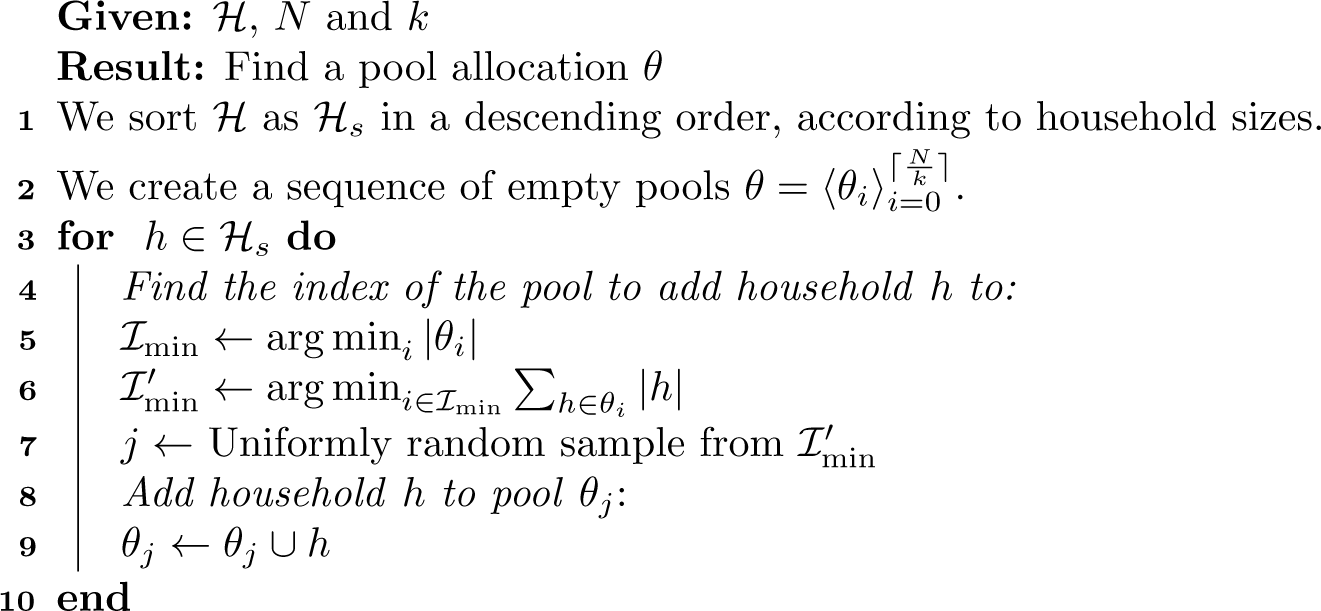

### 2.2 Individual-based model

We use the *STRIDE* individual-based model to simulate households and the social interactions that take place within and between these households [24]. *STRIDE* has been used to reproduce the Belgian COVID-19 epidemic and to evaluate different strategies to gradually exit the lock-down [19]. This model, to which we will refer as *COVID-STRIDE*, was able to closely match the data that was observed during the Belgian epidemic (i.e., hospital admissions, serial sero-prevalence and serial prevalence) [19, 25, 26]). We present a schematic overview of the infection and disease phases individuals can experience in the COVID-STRIDE model in Figure 1. In this study we use the parametrization used by Willem et al., of which we give an overview in Table 1 [19]. This parametrization produces a generation interval of 5.16 days, a doubling time of 3 days and a basic reproduction number of 3.41 (confidence intervals in [19]).

**Table 1:** Disease characteristics and references. Transition parameters are discretized to the modelling time step of 1 day.

| Parameter | Model value | References |
| --- | --- | --- |
| Incubation period | 5-7 days (uniform) | [27, 28] |
| Symptomatic period | 4-6 days (uniform) | [28] |
| Infectious period | 5-7 days (uniform) | [28] |
| Pre-symptomatic infectious period | 2-3 days (uniform) | [28] |
| Hospitalization probability (0-18y) | 0.05 | [29, 30, 25] |
| Hospitalization probability (19-59y) | 0.03 | [29, 30, 25] |
| Hospitalization probability (60-79y) | 0.12 | [29, 30, 25] |
| Hospitalization probability (+80y) | 0.60 | [29, 30, 25] |
| Hospitalization delay (0-18y) | 3 days | [29, 30, 25] |
| Hospitalization delay (19-59y) | 7 days | [29, 30, 25] |
| Hospitalization delay (60-79y) | 7 days | [29, 30, 25] |
| Hospitalization delay (+80y) | 6 days | [29, 30, 25] |
| Infectiousness asymptomatic case | 50% | [31] |
| Proportion of symptomatic cases | Age-dependent | [19, 32] |
| Children’s susceptibility | 50% | [19] |

**Figure 1:**
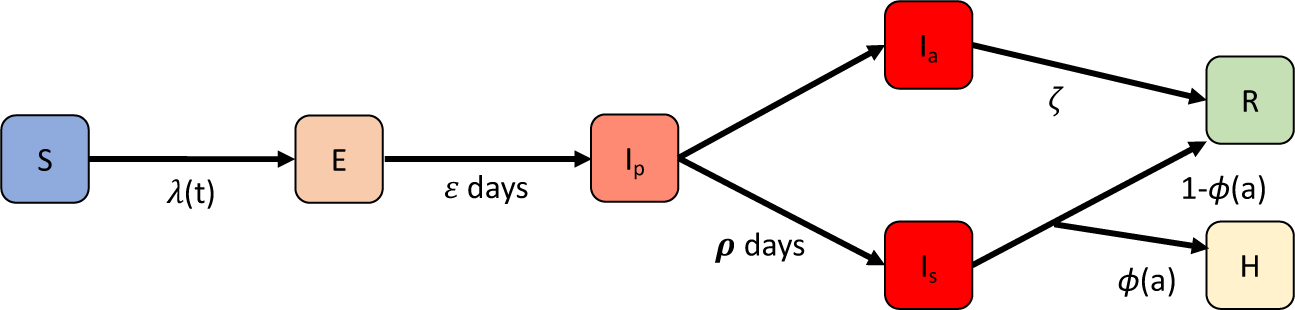
We consider that an individual goes through different phases of infection/disease, which is represented by a SEIR-like state machine. A susceptible individual (S) can become infected, given a time-dependent infection probability *λ* (*t*). This probability depends on the transmission potential of the virus and the social contact behaviour, which due to contact reduction policies is time-dependent. When infected, the individual becomes exposed (*E*). Once exposed (*E*), an individual goes through an incubation time of *ε* days, after which the individual becomes infectious prior to symptom development (*I*_*p*_). A pre-symptomatic infected individual (*I*_*p*_) will either become asymptomatic (*I*_*a*_), symptomatic with symptoms (*I*_*s*_), after a period of *ρ* days. When asymptomatic (*I*_*a*_) the individual will remain infectious for *ζ* days after which he/she recovers (*R*). When severely symptomatic, the individual will be hospitalized with an age-dependent probability *ϕ* (*a*) or recover without the need for hospitalisation.

We extend the *STRIDE* model to incorporate universal testing as described in Section 2.3. The source code of the *STRIDE* model is available as free (GPLv3) software (Section 9).

### 2.3 Pooled-households universal testing

We assume to have a fixed set of PCR tests available per day, *T*_*d*_. Each PCR test is used to test one pool of individuals, *θ*_*j*_, as defined Section 2.1. As we assume that pools have a fixed capacity *k*, this means that we can test up to *k·T*_*d*_ individuals per day. We define one *testing sweep*, as the period (i.e., number of days) it takes to test the entire population, given the ability to test *k ·T*_*d*_ individuals per day. We define universal testing as a repetition of *testing sweeps* to test the entire population.

By testing household members together, our approach facilitates two isolation strategies: *pooled isolation*, i.e., isolating all members of the infected pools, and *individual isolation*, i.e., performing an additional testing step to identify which households are effectively positive. In the case of *pooled isolation*, we isolate each individual for 7 days [33]. In the case of *individual isolation*, we keep the individuals in the pool isolated until it is clear, through individual testing, which individuals are positive. These positive individuals are then kept in isolation until they have been isolated for a total of 7 days [33].

To plan which households will be tested at which day of a testing sweep, we perform two steps. Firstly, we use the allocation algorithm to appoint households to pools (see Section 2.1). To improve the feasibility with respect to the logistics of the testing procedure, we construct a sequence of pools per province (i.e., an administrative region in Belgium, NUTS-2 level^3^). Secondly, for each of the provinces, we assign the obtained pools to the different days of the sweep, such that on each day of the testing sweep, a portion of the province is tested that is proportional to the province’s population size. This assignment will be used for each of the testing sweeps (i.e., repetitions) that will be conducted.

Once this planning has been established, the *COVID-STRIDE* model will, for each simulated day that universal testing is enabled, iterate over all pools and determine whether a pool tests positive. In order to permit a realistic modelling framework for universal testing, we consider the following parameters: testing compliance *c*_*t*_ (i.e., the fraction of households that will cooperate and allow for the test to be performed), isolation compliance *c*_*i*_ (i.e., the fraction of households that will, when asked to self-isolate, comply to the request), the number of days it takes to get the results from the test *d*_*t*_ and the false negative rate of the PCR test FNR_PCR_. When evaluating the *individual isolation* strategy, we consider the same false negative rate for individual testing. For each pool *θ*_*i*_, we first determine which of the households in the pool comply to testing, using a Bernoulli experiment *B*ern(*c*_*t*_) for each household in *θ*_*i*_. We then determine whether one of the compliant households has an individual that has been infected for at least PCR_delay_ days, such that this infection can be picked up by a PCR test. This is determined based on the infection status of the individuals that make up the household, which is encoded in the state of the individual-based model. We choose a delay of 2 days (i.e., PCR_delay_ = 2), informed by the time onset of the pre-symptomatic infectious period (see Table 1), and in line with earlier work [13]. When positive, we determine with a Bernoulli test *B*ern(1 *−* FNR_PCR_) whether the PCR test will detect it. When detected, we isolate either the infected individuals (individual isolation strategy), or all of the households in pool *θ*_*i*_, with a per-household compliance that is determined by a Bernoulli test *B*ern(*c*_*i*_). We conduct our experiments by executing 5 stochastic trajectories of the *COVID-STRIDE* model, for each combination of the parameters we investigate, to assess the stochastic variation of the model.

## 3 Results

We conduct a series of experiments to investigate the proposed universal testing procedure under different assumptions. All of the experiments consider the Belgian COVID-19 epidemic that was fitted to data observed during the epidemic [19]. We assume that individuals are less infectious when asymptomatic (50% reduction in infectiousness) [31]. Furthermore, we assume that 7% of children (i.e., individuals aged 0-19 year) is symptomatic [19]. We use the transmission model from Willem et al., that was calibrated assuming that children are only half as susceptible compared to adults [19]. Given these assumptions, we reproduce the history of the lock-down, to obtain an age-specific immunity profile. To reproduce the lock-down scenario, such that it fits the data that was observed during the Belgian COVID-19 epidemic (i.e., hospitalization and sero-prevelance data) [19], we impose a 75% workplace contact reduction and a 90% leisure contact reduction. Furthermore, we assume that schools (including tertiary education) were closed during lock-down.

To evaluate the household-pooled universal testing procedure, we consider a relieve of the lock-down, at which time we start performing testing sweeps (i.e., repetitions, defined in Section 2.3). We consider this relieve at two time-points: the first of May, with about 50000 active infections^4^, and the first of July, with about 1000 active infections. These two starting points allow us to differentiate between an epidemic with a downward trend that still has a large number of active cases, and an epidemic that is under control, yet prone to experience outbreaks. When the lock-down ends, we assume that the work contact reduction is 50% (compared to 75% during lockdown) [34] and the leisure contact reduction is 70% (compared to 90% during lockdown). Additionally, we open all schools, except for tertiary education. Concerning school openings, we disregard any holidays, to be time-invariant, and thus keep our experiments as generic as possible. Note that all contact reductions, both during and after lockdown are relative to pre-lockdown observations that were obtained in a social contact survey [35].

To explore the efficiency and robustness of our universal testing approach, we consider different values for the universal testing model parameters. Firstly, we consider a pool size *k* ∈ {16, 32}. The pool size impacts the isolation policy, as it dictates the amount of households that need to be isolated when pool isolation is used. We consider the false negative rate of the PCR test, FNR_PCR_ *∈* {0.1, 0.05}. This choice is motivated by the study of Yelin et al. that reports a false negative rate of 10% when using PCR pools up to size 32, yet we also consider a lower false negative rate of 5%, regarding the prospect of improved test pool protocols, or the use of smaller pool sizes (e.g., *k* = 16). Secondly, we consider the availability of PCR tests per day *T*_*d*_ ∈ {25k, 50k}, ^5^ as motivated in the introduction section and further discussed in Section 7. Thirdly, we consider household compliance with respect to testing, *c*_*t*_ ∈ {0.8, 0.9}, and isolation, *i*_*t*_ ∈ {0.8, 0.9}, to evaluate the robustness of our testing framework, when cooperation with the universal testing policy is imperfect. Finally, we assume that the pooled PCR test results can be reported to individuals within 1 day (i.e., *d*_*t*_ = 1). We assume that a one day turn-around time is reasonable, when the extraction of samples and the PCR testing of these samples can be organized in a decentralized fashion. This way, tests can be carefully planned, rather than that they need to be performed on demand (e.g., in the case of contact tracing), and can be collected and analysed in the same location. In Section 5, we discuss these logistic considerations in more detail, to further motivate this reasoning. Furthermore, we conduct a sensitivity analysis considering *d*_*t*_ = {1, 2, 3, 4} in Appendix C. We present an overview of the different parameter values in Table 2.

**Table 2:** Overview of the model parameters related to the universal testing framework.

| Parameter | Values |
| --- | --- |
| Pool size ($k$) | 16, 32 |
| PCR false negative rate ($\text{FNR}_{\text{PCR}}$) | 5%, 10% |
| PCR tests per day ($T_d$) | 25k, 50k |
| Isolation compliance ($c_i$) | 80%, 90% |
| Test compliance ($c_t$) | 80%, 90% |
| Reporting delay ($d_t$) | 1 day |

To investigate the impact of our household-pooled universal testing approach, we obtain 5 stochastic trajectories of the *COVID-STRIDE* model, for each combination of these parameters. When analysing these results, we observe that the main trends of the model results are due to the amount of days one sweep takes to complete. As this is determined by the pool size *k* and the number of PCR tests that are available per day *T*_*d*_, we show these trends by aggregating all model results per combination of ⟨*k, T*_*d*_⟩, for each of the considered false negative rates FNR_PCR_. In Figure 2, we show the trends for both starting points and a false negative rate FNR_PCR_ = 0.1 (i.e., the least optimistic of the considered false negative rates), and we follow the pool-isolation strategy, where we isolate all individuals that are part of an infected pool. This figure demonstrates that, in our simulations, we can only achieve proper control of the epidemic, i.e., when *k* = 32 and *T*_*d*_ = 50000. This parameter combination allows for the highest number of individuals to be tested per day (i.e., the whole population in one week), which shows a rapid decrease of the number of infections. Similar trends were observed when the false negative rate is 5%, and these results are shown in Appendix A. To demonstrate the performance gradient between weekly testing and bi-weekly testing, we also show results for *k* = 32 and *T*_*d*_ ∈ {25k, 30k, 35k, 40k, 45k, 50k} in Appendix E.

**Figure 2:**
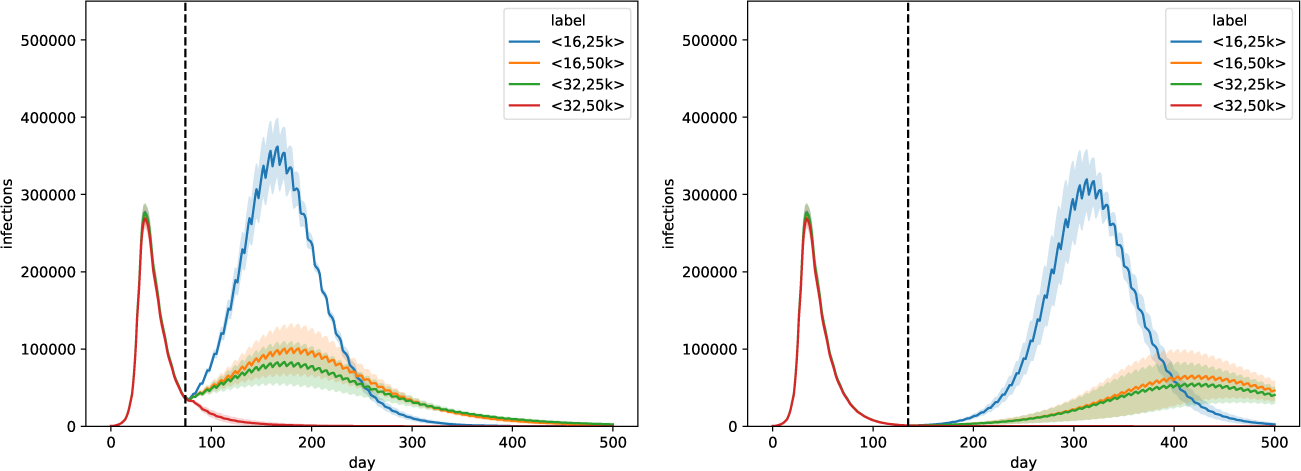
Trends for pool isolation. Trends for all combinations of parameters ⟨*k, T*_*d*_⟩, for FNR_PCR_ = 0.1. Universal testing starts at the first of May (left panel) and the first of July (right panel). We follow the pool isolation strategy, where we isolate all individuals that are part of an infected pool. The curves show a line that depicts the average over the trajectories of the result aggregations and a shaded area that depicts the standard deviation.

We compare these results to the individual isolation strategy in Figure 3, where we determine which of the individuals that belong to the positive pool are actually positive. Overall, the results with pool isolation and individual isolation are similar. However, due to the additional test that is required for each of the individuals that belong to a positive pool, and the false negative rate that is again associated with it, the performance of individual testing is lower. We observed similar trends for the 5% false negative rate, and these results are shown in Appendix A.

**Figure 3:**
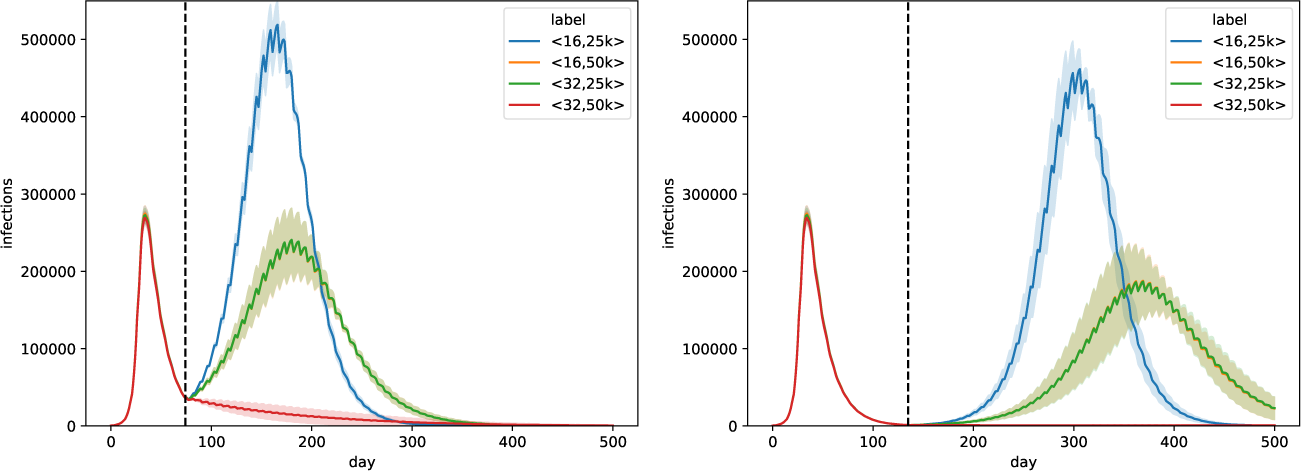
Trends for individual isolation. Trends for all combinations of parameters ⟨*k, T*_*d*_ ⟩, for FNR_PCR_ = 0.1. Universal testing starts at the first of May (left panel) and the first of July (right panel). We follow the individual isolation strategy, where we identify the infected individuals in positive pool. The curves show a line that depicts the average over the trajectories of the result aggregations and a shaded area that depicts the standard deviation. Note that in this figure, the orange and green curve overlap.

To demonstrate the influence of test and isolation compliance on the decrease in infections, we show box-plots that depict the number of infections at three different time points (i.e., 90 days, 180 days and 270 days after the start of the universal testing procedure), for the experiment when the lock-down ends on the first of July. We show the results for the two isolation strategies, respectively pool isolation in Figure 4 and individual isolation in Figure 5, for the weekly universal testing procedure (i.e., *k* = 32 and *T*_*d*_ = 50000). In Figure 4 and Figure 5 we show results for a false negative rate of 10%. Similar trends were observed for the other (lower) false negative rate, and these results are shown in Appendix A. These results demonstrate that we converge to zero cases for both isolation strategies, except when individual isolation is used, and the compliance of both test and isolation is below 80%. Due to the additional test that is required for each of the individuals that belong to a positive pool, and the false negative rate that is again associated with it, individual isolation exhibits a lower convergence rate. Another important consideration here, is the difference in compliance between the two isolation strategies, as we argue that individuals will more likely comply when they know that they are infected. We note that, when compliance is higher, the effect of individual isolation is less pronounced. Note that, under the leisure and work contact reductions that we assume, the cases converge to zero over a time span that covers many months. In Section 4, we show that the number of cases can drop more quickly, when the contact reduction is more pronounced.

**Figure 4:**
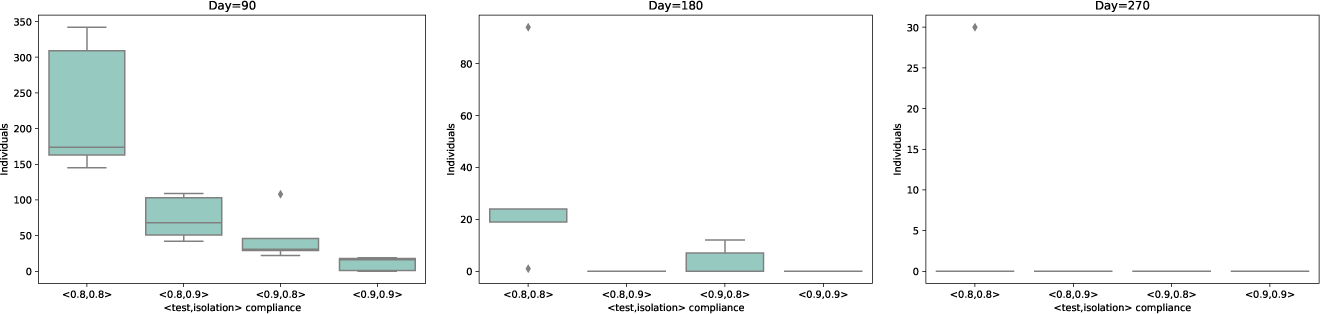
Distribution for pool isolation. Distribution of the number of infections for the experiment when the lock-down ends on the first of July, in different scenarios of compliance for testing and isolation. We show the number of infections at three different time points. i.e., 90 days (left panel), 180 days (middle panel) and 270 days (right panel) after the start of the universal testing procedure. These results consider a weekly universal testing procedure (i.e., *k* = 32 and *T*_*d*_ = 50000) and a FNR_PCR_ = 0.1, where the isolation strategy is pool isolation. Each box represents a combination of test and isolation compliance.

**Figure 5:**
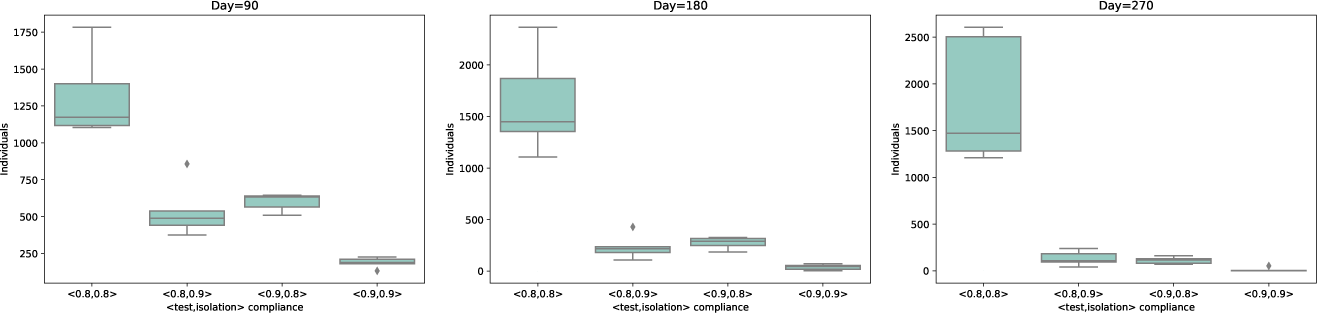
Distribution for pool isolation. Distribution of the number of infections for the experiment when the lock-down ends on the first of July, in different scenarios of compliance for testing and isolation. We show the number of infections at three different time points. i.e., 90 days (left panel), 180 days (middle panel) and 270 days (right panel) after the start of the universal testing procedure. These results consider a weekly universal testing procedure (i.e., *k* = 32 and *T*_*d*_ = 50000) and a FNR_PCR_ = 0.1, where the isolation strategy is individual isolation. Each box represents a combination of test and isolation compliance.

In the previous experiments, we consider a fixed leisure (post lock-down) contact reduction of 70%. To assess the amount of contacts that can be allowed when performing weekly universal testing, we investigate different leisure contact reductions, while keeping the work contact reduction fixed to 50%. To this end, in Figure 6, we show the simulation results for a leisure contact reduction of 50%, 60% and 70%, for both isolation strategies. To extend this setting to allow travel, we consider importing *n* cases per day, i.e., *n* individuals that were infected during a travel and return home. We show these results for respectively *n* = 10 and *n* = 50 in Figure 7 and 8. In Figure 6, Figure 7 and Figure 8 we show results for a false negative rate of 10%, and similar trends were observed for the 5% false negative rate (Appendix B). These results show, in simulation, that control of the epidemic can be achieved for leisure contact reductions up to 60%, when pool isolation is used. With individual isolation, the performance is lower, due to the additional test that needs to be performed, and the false negative rate that is associated with it. The pool isolation strategy can keep the epidemic at a reasonable level for a contact reduction of 50%, yet given this incidence level observed, this isolation strategy would require the isolation of many uninfected individuals, potentially impacting community compliance.

**Figure 6:**
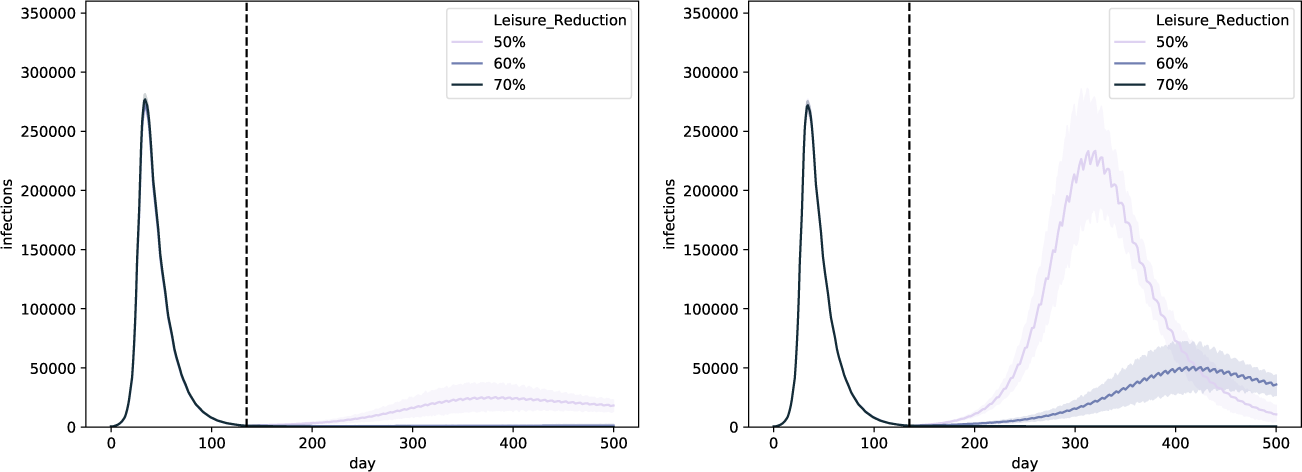
Trends for different leisure contact reductions, when performing weekly universal testing. We assume that universal testing starts on the first of July and that FNR_PCR_ = 0.1. We consider both isolation strategies: pool isolation (left panel) and individual isolation (right panel). The curves show a line that depicts the average over the trajectories of the result aggregations and a shaded area that depicts the standard deviation.

**Figure 7:**
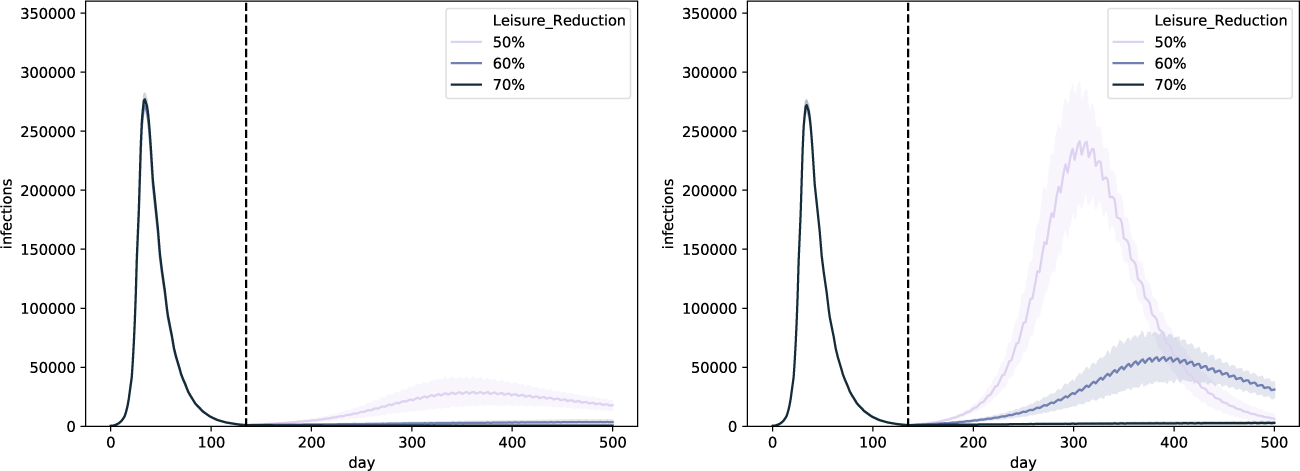
Trends for different leisure contact reductions, when performing weekly universal testing, and importing 10 cases per day. We assume that universal testing starts on the first of July and that FNR_PCR_ = 0.1. We consider both isolation strategies: pool isolation (left panel) and individual isolation (right panel). The curves show a line that depicts the average over the trajectories of the result aggregations and a shaded area that depicts the standard deviation.

**Figure 8:**
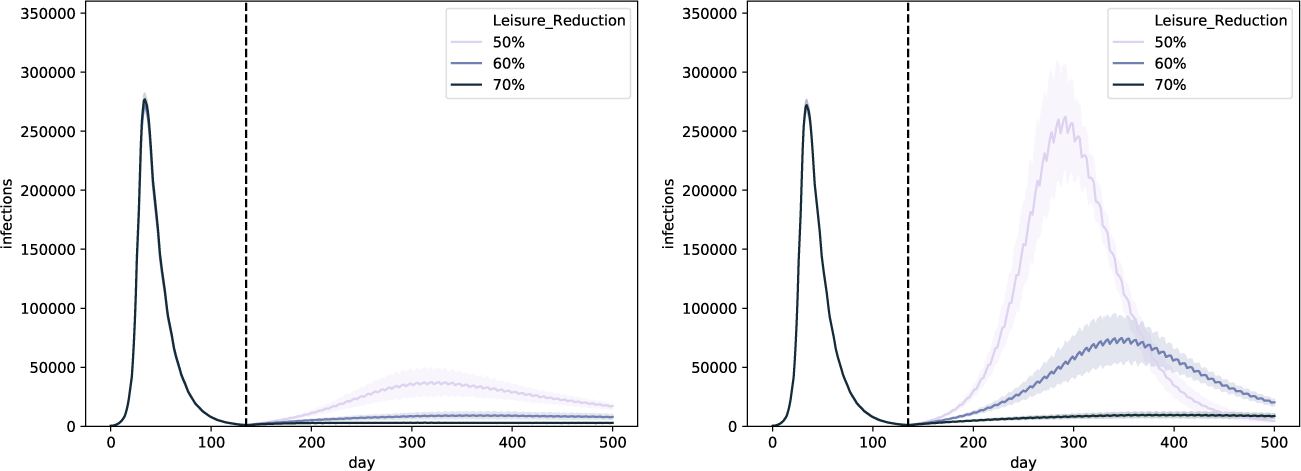
Trends for different leisure contact reductions, when performing weekly universal testing, and importing 50 cases per day. We assume that universal testing starts on the first of July and that FNR_PCR_ = 0.1. We consider both isolation strategies: pool isolation (left panel) and individual isolation (right panel). The curves show a line that depicts the average over the trajectories of the result aggregations and a shaded area that depicts the standard deviation.

Note that we choose the same false negative rate for the pool test and the individual test, which is a conservative assumption, given that the individual test can be performed using the RNA extract that was established during the pool test. As the false negative rate compounds both errors with respect to sample collection and the sensitivity of the PCR test [36], it is reasonable to assume that the follow-up PCR test will have a lower false negative rate, as the virus was already extracted successfully from the individuals that are part of a positive pool. It is however challenging to choose a realistic reduction of the false negative rate for the additional test, as this is complicated when a pool is made up out of multiple positive samples and each of these samples can independently result in an error. Our experiments in Appendix A show that a lower false negative rate results in improved performance when individual isolation is applied. Therefore, we believe follow-up research is warranted to obtain an improved estimate of the false negative rate of individual tests that use the RNA samples obtained from a pooled PCR test.

Our results also indicate that the importation of cases has an impact, which is more pronounced for lower leisure contact reductions, and when individual isolation is used. Overall, when the contact reduction is 70%, both isolation strategies are capable to control the epidemic.

In these experiments, we assume that when isolation is imposed, individuals are able to isolate from household members as well. When individuals are aware of their infection status, as is the case when individual isolation is applied, this assumption is reasonable and in line with earlier work [19]. However, we argue that this is less straightforward to accomplish in the case of pool isolation. Therefore, we challenge this assumption in Appendix C. These experiments shows that the model trends are robust to this effect when the contact reduction is 70%. However this effect becomes pronounced when the number of contacts is increased (50%,60%). This highlights the importance of social distancing in the household, when a household member has been found positive.

To asses the impact of the PCR test reporting delay *d*_*t*_, we conduct a sensitivity analysis considering *d*_*t*_ = {1, 2, 3, 4} in Appendix D. On the one hand, this sensitivity analysis shows that the pool isolation strategy is quite robust with respect to waiting times up to four days (Figure 22). On the other hand, our experiments show that for the individual isolation, the effect of this delay is much more pronounced and even dramatic for *d*_*t*_ ≥ 3 (Figure 22). The reason for this strong effect is that, while individuals remain unaware of their infection status, it is likely that they will infect their household members. This effect is reduced when pool isolation is used, as the household members are part of the same pool and will thus be isolated as well. Based on this observation, we hypothesised that the use of *household isolation* (i.e., individual testing is used to determine the infection status of individuals and all households that contain infected individuals are isolated as a whole) could reduce this effect, which is confirmed experimentally in Figure 23 (Appendix D).

## 4 Universal testing and “the hammer”

In Section 3, the cases converge to zero over a time span that covers several months, given the leisure and work contact reductions that we assume. Here, we investigate the effect of weekly universal testing when it is conducted under contact reductions that are the same as during the lock-down that took place in Belgium during the first wave of the epidemic, i.e., a 75% workplace contact reduction and a 90% leisure contact reduction, also referred to as “the hammer”. In Figure 9, our simulation results show that the cases drop to zero quickly, even when considering lower levels of isolation and testing compliance.

**Figure 9:**
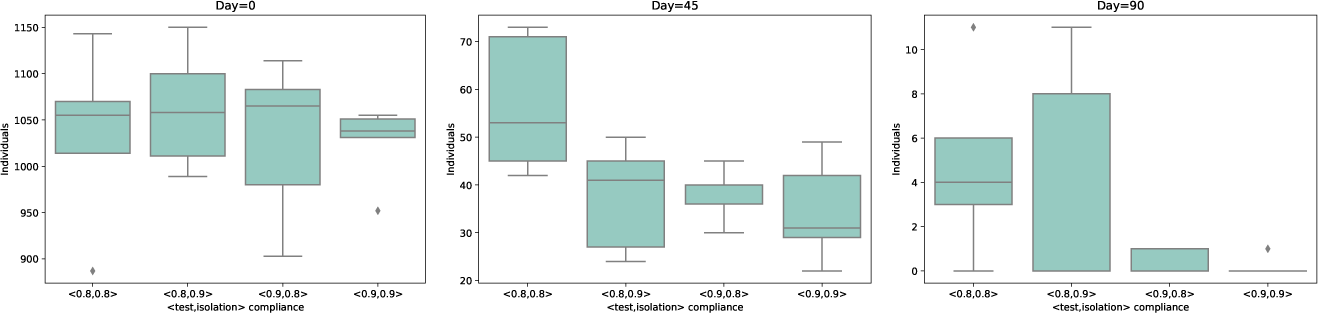
Distribution of the number of infections for the experiment when the lock-down restrictions are continued. We show different scenarios of compliance for testing and isolation. We show the number of infections at three different time points. i.e., 0 days (left panel), 45 days (middle panel) and 90 days (right panel) after the start of the universal testing procedure. These results consider a weekly universal testing procedure (i.e., *k* = 32 and *T*_*d*_ = 50000) and a FNR_PCR_ = 0.1, where the isolation strategy is individual isolation. Each box represents a combination of test and isolation compliance. Note that 135 days after the start of the universal testing procedure (not shown in this figure), the number of cases dropped to zero, for all the compliance scenarios.

While resistance towards lock-down measures is growing in many countries throughout the world, our intention is not to promote them here. Yet, this investigation does show that a (fast) local eradication might be feasible when performing a weekly universal testing procedure. Furthermore, this highlights the potential of universal testing strategies as a mitigation mechanism to other (future) emerging infectious diseases, that warrants further investigation.

## 5 Logistics of weekly universal testing

The time it takes to test the entire population (i.e., the sweep time), is directly proportional to the frequency at which each individual should be tested. Our experiments show that a sweep time of 1 week, of a population of 11 million individuals (i.e., using a fixed pool size *k* = 32 and a constant amount of tests *T*_*d*_ = 50k), yields the most promising results. We acknowledge that testing each individual on a weekly basis, requires a significant effort from the community. Yet, our simulation results do show that these actions may result in the control of the epidemic, even when much of the contact reductions are relieved, which might convince individuals that these measures are worth the effort. Notwithstanding, in order to make such a policy successful in a public health context, it is important that the societal awareness and support of these policies is stimulated, via prompt governmental communication.

Next to the efforts required by the population to get tested on a weekly basis, there is also a significant logistic challenge to obtain samples and ensure they can be tested in a reasonable amount of time. We discern four main considerations with respect to the logistics related to universal testing. Firstly, to enable a suitable geographical planning of the sample extraction and PCR testing, we argue that a decentralized approach will be most suitable. In our experiments, we already establish the test planning on a provincial level. In these provinces, a further division of this allocation towards regions that have PCR testing capabilities is possible, thereby facilitating a decentralized approach. Secondly, in order to perform sample extractions, a significant number of nursing staff members is required, for which we compute an estimate. For clarity, we express this computation in terms of a 1000 individuals, which can be extrapolated to the whole population in a straightforward way. When we assume that tests are collected over the time of 8 hours per day and every day of the week (including weekends), we need to test 18 individuals per hour, to test this set of 1000 individuals in one week. When we consider the use of a drive-through sample extraction facility, it is reasonable to assume that 1 nurse can test 9 individuals per hour, especially as a significant fraction of these individuals are part of the same household. When we assume that nurses work for 5 days per week and 8 hours per day, we conclude that we need on average 2.8 nurses per thousand individuals. Thirdly, when the samples are extracted, they have to be pooled and PCR-tested using the available PCR-testing infrastructure. In our experiments, we assume that 50k PCR tests are available per day to test the sample pools. This assumption is in line with the objective of the Belgian government to provide 70k PCR tests per day [20]. Furthermore, this leaves 20k PCR tests to identify the individuals that are responsible for the positivity of the pool. Fourthly, it is important that the pools are tested in a reasonable amount of time. While there have been complaints from different countries that the turn-around time of PCR testing in the context of contact tracing was too slow, we believe that the use of universal testing could improve this. This is the case when tests can be planned, rather than performed on demand (in the case of contact tracing), such that the burden on the testing infrastructure is distributed more uniformly. Furthermore, while in the context of contact tracing, sample extraction is commonly conducted by physicians and the extracted samples need to be sent to the PCR testing laboratories, which results in a lot of time spent on transporting the samples. To contrast this, the use of drive-through testing facilities, where sample collection and processing occurs in the same location, reduces the overall turn-around time for testing. Therefore, we believe that careful planning will enable to report the testing results within one day.

We acknowledge that the current testing practice, where a swab is placed in the nose, is quite invasive. In this regard, alternative sampling techniques, such as saliva sampling (recently approved by the FDA [37]), should be considered. As saliva testing is a less invasive testing procedure, it would likely increase testing compliance. Furthermore, the use of saliva sampling could allow individuals to self-collect their samples, which could reduce the complexity of the testing logistics.

## 6 Related work

We take note of several works that relate to our method.

The work by de Wolff et al. also investigate pooling as a faster and more resource-efficient alternative to individual testing and present a method to determine which samples in a pool are positive using as little tests as possible [38]. Although in our work, we mostly focus on the phase of the epidemic where the number of infections is low, this technique could still be used in conjunction with the method that we propose.

The work by Taipale et al. considers a more accessible and cheaper, yet less reliable test, to infer whether individuals are infected [39]. This approach does rely on different chemical reagents than the ones currently kept in storage by governments. Therefore, implementing this method would require a change in the testing procedures, while the method that we propose could in principle be used right away [39], using the already implemented testing procedure and may therefore be utilized without further logistic alterations.

Recently, a platform, Swab-Seq, to perform massively scaled SARS-CoV-2 testing was introduced by Bloom et al. [40]. This technique allows to test thousands of samples simultaneously, by pooling these samples together in a well. This new technique is interesting, yet again, it still needs to be implemented in lab environments, to be used widely. As a side-note, while this scalability is impressive, we believe that a household-pooled testing approach could enable a faster turn-around time. On the one hand, for Swab-Seq to work for universal testing, thousands of samples need to be collected before the well is filled, and as Bloom et al. report, processing a well takes between 12 and 24 hours, resulting in a long overall turn-around time. On the other hand, the sample pooling approach we propose allows the extraction of samples that belong to a certain pool (e.g., *k∈* {16, 32}), after which this pool can be send directly for PCR testing.

Recently, the idea of fast and cheap antibody tests was introduced by Mina et al. ^6^, that aim to test individuals on a daily basis. The prospect of the wide availability of such tests is promising, and we believe that our modelling framework could prove useful to investigate the optimal allocation of such tests.

## 7 Discussion

In this study, we propose a method that renders universal testing feasible, requiring an amount of PCR tests per day, that is currently available, or will be made available to accommodate the expected increase in respiratory infections in autumn [21]. We evaluate this method using the *COVID-STRIDE* individual-based epidemic model and investigate how this universal testing method can be used to control a local SARS-CoV-2 epidemic. This evaluation highlights two important results. Firstly, in simulation, our method for universal testing is able to keep the epidemic under control, when each individual can be tested on a weekly basis, even when many of the social contact restrictions are lifted (i.e., opening of schools, work contacts, leisure contacts). Secondly, in simulation, we show that, with increased social contact reductions and without travel, the universal testing approach, allows for the number of infected individuals to converge to zero, rendering it a possible approach to eradicate SARS-CoV-2.

In this study, we use the *COVID-STRIDE* individual-based model, a mechanistic model that was fitted to data that was recorded during the Belgian SARS-CoV-2 epidemic [19]. The extensive validation in [19] shows that *COVID-STRIDE* allows to accurately model the transmission of SARS-CoV-2 within and among households. Our extension to the *COVID-STRIDE* model, with respect to universal testing, aims to reflect a realistic framework that can model the effect of difficulties and issues in the implementation of the intervention strategy, such as the false negative rate of the PCR test and difference in compliance, both with respect to isolation and testing. Nevertheless, we acknowledge that our epidemiological model is an abstraction of the real world that relies on a series of assumptions, and therefore the results presented in this manuscript should be interpreted with caution.

An important assumption in this work is the false negative rate of the PCR test, when a pool of samples is processed. We consider a range of false negative values (i.e., 5% and 10%), predominantly informed by the recent work of Yelin et al. [18]. This study associates the false negative rate to the pool size (i.e., more samples in the pool leads to a higher false negative rate), and attributes these differences to the effect of the dilution of the sample.

To assess policies in a realistic framework, it is essential to consider difficulties and issues in the implementation of the intervention strategy, as they will be inevitable if the policy is to be implemented. This includes technical properties of the tests, as mentioned above, but also the willingness of individuals to participate. To this end, we validated our policies considering different levels of compliance with respect to testing and isolation. Unsurprisingly, our simulation results show that higher compliance results in a faster and more pronounced drop in the epidemic curve, yet overall our proposed strategy is robust to imperfect compliance. As stated earlier (see Section 5), we stress the importance to gain support from the community, to keep the population motivated to take part in this policy, especially if frequent testing is necessary. Next to compliance with respect to the testing policy, our simulations show that it is important for individuals to comply with the isolation policy that is in place. In this study, we consider two isolation policies: pool isolation and individual isolation. While pool isolation reduces the need for additional tests, we acknowledge that it might be challenging for individuals to adhere to isolation when they are unaware of their infection status. From another perspective, as we aim to move to a low number of infections with our method, the number of pools that need to be isolated will also be low, and the burden of pool isolation might be acceptable. From a logistic perspective, individual isolation seems feasible when the incidence is low and the number of tests necessary to determine which individuals are positive is limited. For the Belgian case, the government’s objective is to have 70k PCR tests available by autumn of 2020. This would allow to test the population on a weekly basis (i.e., using 50k PCR tests and a pool size of 32), and have 20k PCR tests available to detect the infected individuals in the positive pools. We argue that individual isolation of cases would be beneficial with respect to isolation compliance. Analogously, the use of pool isolation could prove beneficial when the incidence is high (see left panel of Figure 2 in Section 3), and the number of individuals of positive pools exceeds the number of available tests. This intervention strategy could be used to avert or shorten future lock-downs, where the isolation of pooled individuals is less burdensome compared to shutting down society in its entirety. Another option to reduce the number of individuals that need to be isolated when using pool isolation, is by adding an additional testing step to determine which household is positive. In contrast to individual isolation, this can be done by combing all samples that belong to the same household and test this pool of household members using a single PCR test. By pooling households members, we can determine which of the household(s) cause the positive test, and isolate this/these household(s) for 7 days. This way, the excessive number of additional tests is reduced to the number of households in the pool, which is a quantity we aim to minimize with our allocation algorithm (Section 2.1).

Next, we discuss the most important practical differences between contact tracing and universal testing. To ensure that contact tracing works, individuals need to be identified as an index case, index cases need to cooperate and share (and remember) their list of contacts, and all this needs to be achieved in as little time as possible [19]. Therefore, it might be challenging to fully control SARS-CoV-2 epidemics through the use of contact tracing [12, 13, 14]. In contrast, universal testing is logistically challenging, but once these logistics are in place, the process is transparent. An important consideration with respect to universal testing is compliance fatigue and this is an aspect that should be closely monitored. We note that this can be monitored, as the failure rate with respect to testing compliance is a by-product of the universal testing procedure. While we demonstrate the potential of universal testing to control local epidemics, there is another advantage given that the procedure implicitly performs surveillance of the whole population. This means that a good estimate of the actual incidence, and the geographic distribution of this incidence, will be available at any time. This way, emergency signals will be picked up more rapidly, enabling a swift response that might avoid more invasive control measures.

We also show, in simulation, that our method has the potential to eradicate SARS-CoV-2 in a local setting. While in our simulations, we investigate the effect of importing cases (i.e., infected individuals that return from a travel), we limit ourselves to a constant number of daily importation events. The implementation in a larger travel union (e.g. European Economic Area, United states of America) would be more challenging, as different countries have different epidemic states, and the travel flows between these countries should be considered. Yet, when the countries in a travel union coordinate and implement the system of universal testing locally, the eradication of SARS-CoV-2 within the travel union could be conceivable, when importations from outside the travel union are carefully monitored (e.g., testing on the airports).

The household-to-pool allocation algorithm that we introduce in Section 2.1 is guided by a simple heuristic that results in an optimal allocation, i.e., to adequately fill up the pools and to minimize the difference in number of households between pools (Section 2.1, Equation 4). However, there are some border cases, where using the heuristic could result in an allocation that exceeds the capacity of some of the pools. We argue that such border cases are unlikely to occur in a population that is large enough (i.e., it thus involves a large number of bins) and has a realistic distribution of household sizes, and for pools that have sufficient capacity. The intuition behind this is that, as we add households ordered by their size, first the largest households that are most likely to generate an overflow are assigned. Therefore, as long as there is a sufficient number of smaller households, which is expected and observed in our household dataset (see Figure 10), these households will act as padding to fill up the pools. We were unable to come up with realistic counter-examples, and in our experiments, no overflows were detected.

**Figure 10:**
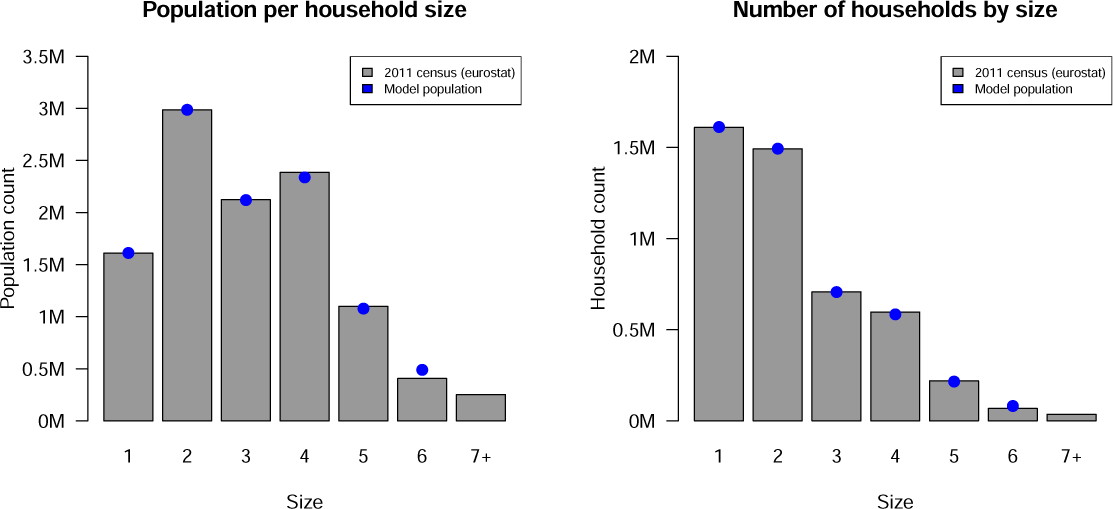
Population size per household size and number of households per size: Belgian 2011 census and model population. Numbers are expressed in million (M). Figure from Willem et al. [19].

One point of concern is that recently recovered individuals could still carry virus particles and render a PCR test positive. While this is not a problem when individual isolation is used, as these individuals can be excluded from the future pool, this could result in false positives when pool isolation is used. While we argue that this artefact is less likely to occur due to sample dilution [18], this warrants further investigation. We note that, if problematic, this issue could be mitigated by pooling households to determine which of the households was infected, as we discussed earlier.

We show in our simulation experiments that the impact of leisure contact reduction has a significant impact on the effectiveness of the weekly universal testing approach. To this end, we conduct a sensitivity analysis considering different levels of leisure contact reduction, yet it remains to be decided how such contact reductions can be translated to a public health context. In this regard, the use of social bubbles of different constellations could be investigated [19], but also the effect of additional hygienic measures (e.g., mouth masks) to reduce the risk of certain contacts could be assessed [41, 42].

Finally, we note that this study was conducted in the context of the Belgian SARS-CoV-2 epidemic, and the reader should be careful to extrapolate these findings to other epidemic contexts. Nevertheless, when the population structure (e.g., household distribution) and size are similar, we believe that the methods presented in this work can be applied to administrative regions of different countries. Furthermore, in this study, we assume 70k PCR tests per day for a population of 11 million (50k PCR tests for weekly testing, and 20k PCR tests to test the individuals in the positive pools), which implies that every day a number of tests is available that corresponds to 0.68% of the population. This number of tests is a reasonable assumption, to which many countries already aspire to anticipate the coming autumn and winter of 2020 [20, 21]. While the logistic requirements to establish weekly universal testing are challenging, we argue and motivate (Section 5) that a decentralized approach is feasible. Furthermore, testing methods such as saliva testing could further reduce the threshold to facilitate universal testing. Acknowledging these complications, we demonstrate that the benefits of universal testing are significant and can result in an improved control and surveillance of the epidemic, resulting in an increase in leisure contacts and an overall societal and economic relaxation.

## 8 Conclusion

In this work, we present a new method to approach universal testing, where we combine households to form sample pools, such that with the available testing capacity, it is possible to test the whole Belgian population on a weekly basis. This weekly universal testing approach allows to isolate pools (when the incidence is high) or to identify and isolate the individuals that are actually positive, using a follow up test (when the incidence is low). On the one hand, such a universal testing approach presents several logistic challenges, which we discuss and for which we formulate a logistic framework. On the other hand, we show in an individual-based model that this mitigation strategy allows an increase in the number of contacts (e.g., work, leisure, schools) and is robust with respect to the importation of cases via travel. We conclude that weekly universal testing could prove an additional strategy to control SARS-CoV-2 outbreaks. Furthermore, we show that the use of universal testing in combination with stringent contact reductions could be considered as a strategy to eradicate the virus. To allow for weekly universal testing to be used in practice, our robustness analyses show that both compliance support from the community and an adequate organisation of sampling logistics is crucial.

## Data Availability

The source code for the simulation model is freely (GPLv3) available on GitHub (universal branch): https://github.com/lwillem/stride.
This work generates no new data, except for simulation results, which can be generated from the simulation model source code.
All of the data used in our simulation model is mentioned in the reference section.

## 9 Availability of data and materials

The source code for the simulation model is freely (GPLv3) available on GitHub (universal branch): https://github.com/lwillem/stride. This work generates no new data, except for simulation results, which can be generated from the simulation model source code. All of the data used in our simulation model is mentioned in the reference section.

## 11 Acknowledgements

The authors are grateful for access to the data from the Belgian Scientific Institute for Public Health, Sciensano.

## A Model results for FNR_PCR_ = 0.05

**Figure 11:**
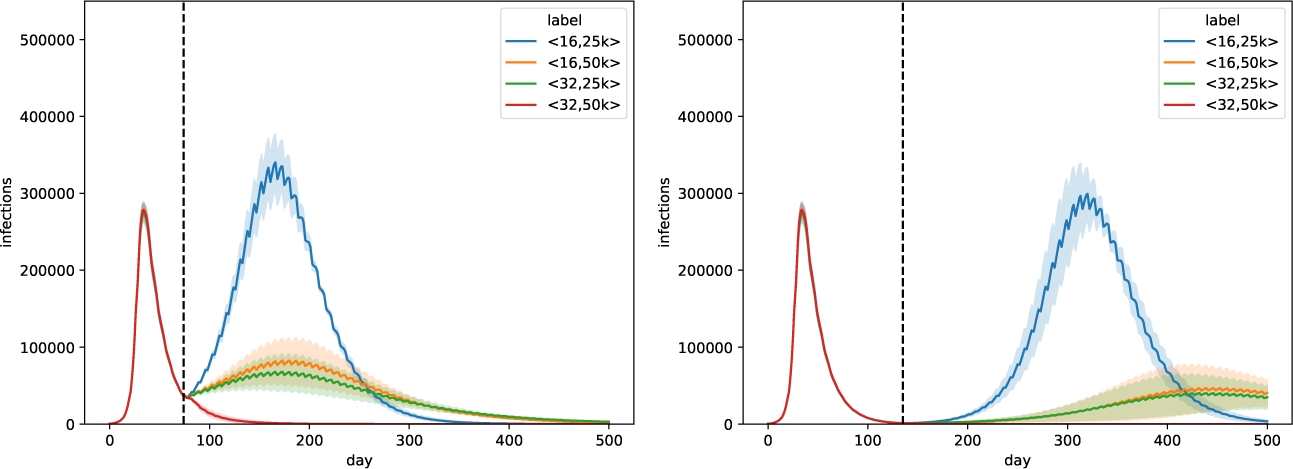
Trends for pool isolation. Simulated epidemic curves for all combinations of parameters ⟨*k, T*_*d*_ ⟩, for FNR_PCR_ = 0.05. Universal testing starts at the first of May (left panel) and the first of July (right panel). We follow the isolation strategy where we isolate all individuals that are part of an infected pool. The curves show a line that depicts the average over the trajectories of the result aggregations and a shaded area that depicts the standard deviation.

**Figure 12:**
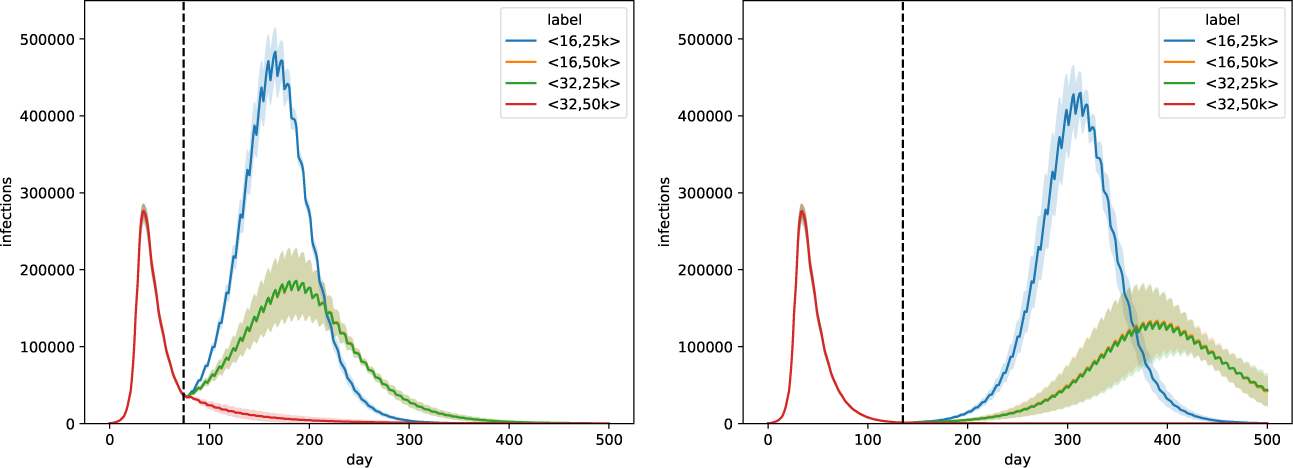
Trends for individual isolation. Simulated epidemic curves for all combinations of parameters ⟨*k, T*_*d*_ ⟩, for FNR_PCR_ = 0.05. Universal testing starts at the first of May (left panel) and the first of July (right panel). We follow the isolation strategy where we identify the infected individuals in the positive pool. The curves show a line that depicts the average over the trajectories of the result aggregations and a shaded area that depicts the standard deviation.

**Figure 13:**
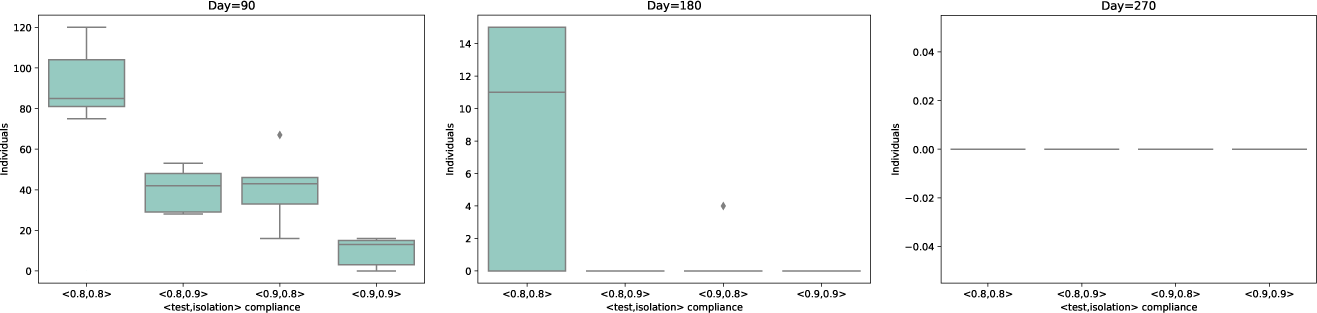
Distribution for pool isolation. Distribution of the number of infections for the experiment when the lock-down end on the first of July, in different scenarios of compliance for testing and isolation. We show the number of infections at three different time points. i.e., 90 days (left panel), 180 days (middle panel) and 270 days (right panel) after the start of the universal testing procedure. These results consider a weekly universal testing procedure (i.e., *k* = 32 and *T*_*d*_ = 50000) and a FNR_PCR_ = 0.05, where the isolation strategy is pool isolation. Each box represents a combination of test and isolation compliance.

**Figure 14:**
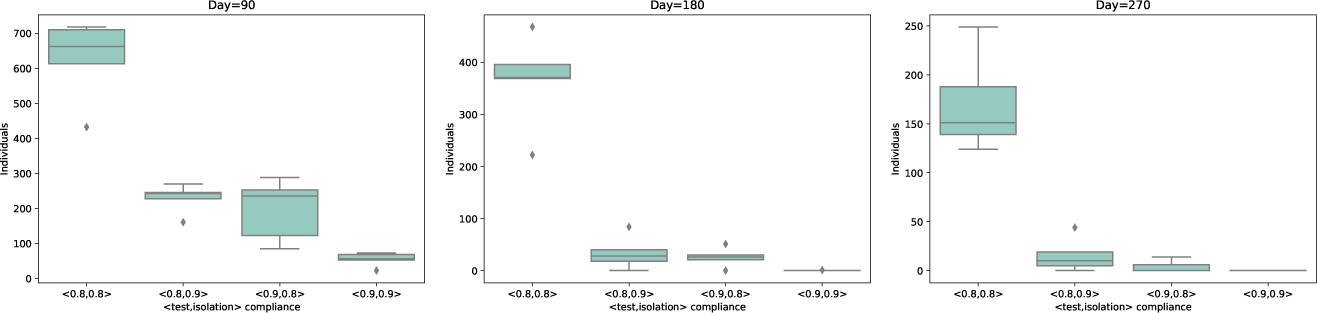
Distribution for individual isolation. Distribution of the number of infections for the experiment when the lock-down end on the first of July, in different scenarios of compliance for testing and isolation. We show the number of infections at three different time points. i.e., 90 days (left panel), 180 days (middle panel) and 270 days (right panel) after the start of the universal testing procedure. These results consider a weekly universal testing procedure (i.e., *k* = 32 and *T*_*d*_ = 50000) and a FNR_PCR_ = 0.05, where the isolation strategy is individual isolation. Each box represents a combination of test and isolation compliance.

## B Model results for different leisure reductions FNR_PCR_ = 0.05

**Figure 15:**
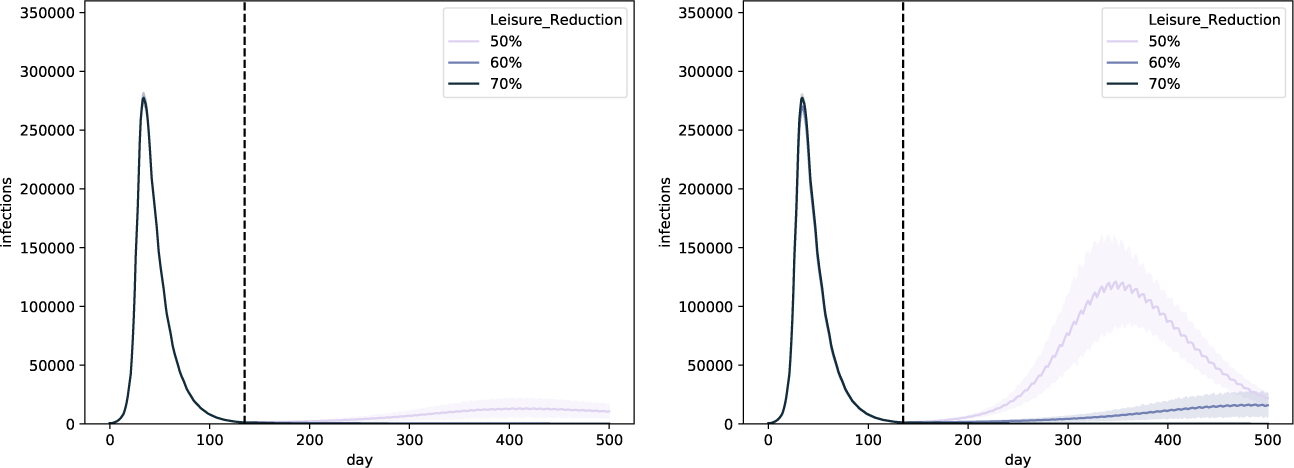
Simulated epidemic curves for different leisure contact reductions, when performing weekly universal testing. We assume that universal testing starts on the first of July and that FNR_PCR_ = 0.05. We consider both isolation strategies: pool isolation (left panel) and individual isolation (right panel). The curves show a line that depicts the average over the trajectories of the result aggregations and a shaded area that depicts the standard deviation.

**Figure 16:**
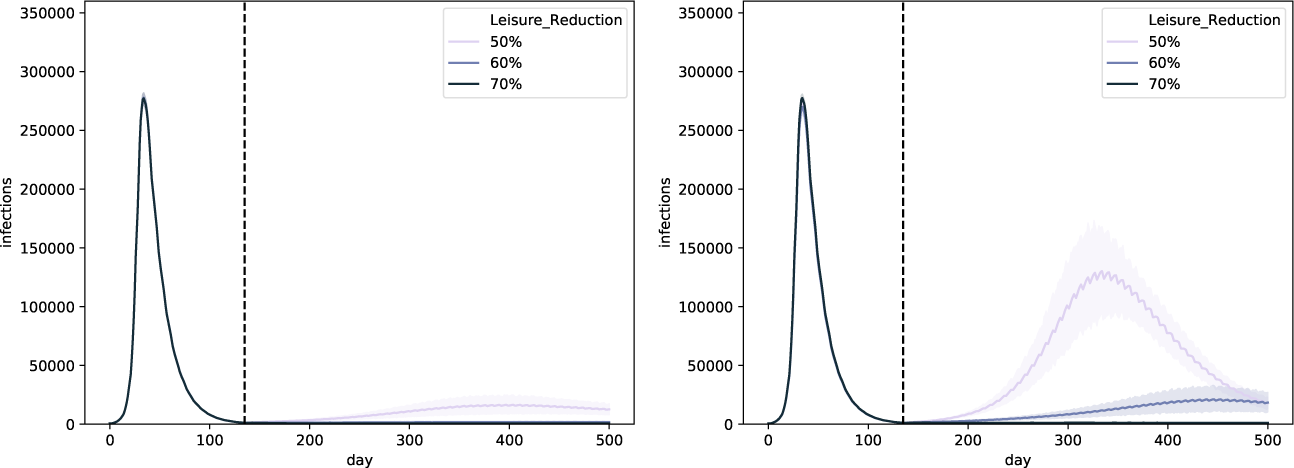
Simulated epidemic curves for different leisure contact reductions, when performing weekly universal testing, and importing 10 cases per day. We assume that universal testing starts on the first of July and that FNR_PCR_ = 0.05. We consider both isolation strategies: pool isolation (left panel) and individual isolation (right panel). The curves show a line that depicts the average over the trajectories of the result aggregations and a shaded area that depicts the standard deviation.

**Figure 17:**
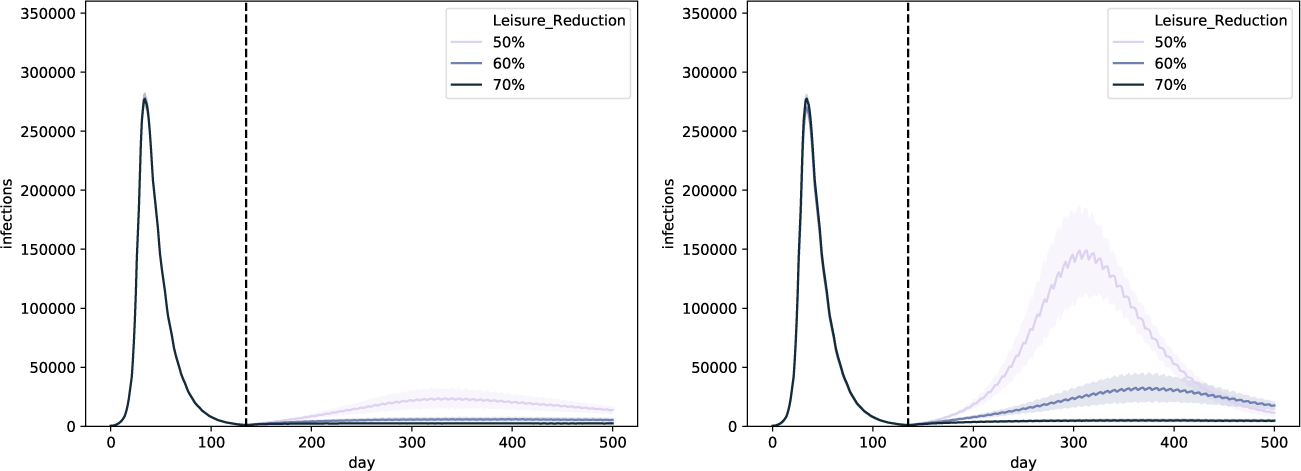
Simulated epidemic curves for different leisure contact reductions, when performing weekly universal testing, and importing 50 cases per day. We assume that universal testing starts on the first of July and that FNR_PCR_ = 0.05. We consider both isolation strategies: pool isolation (left panel) and individual isolation (right panel). The curves show a line that depicts the average over the trajectories of the result aggregations and a shaded area that depicts the standard deviation.

## C Model results for the household isolation challenge

In the main experiments, we assume that when isolation is imposed, individuals are able to isolate from household members as well. When individuals are aware of their infection status, as is the case when individual isolation is applied, this assumption is reasonable and in line with earlier work [19]. However, we argue that this is less straightforward to accomplish in the case of pool isolation. Therefore, in this appendix, we challenge this assumption.

**Figure 18:**
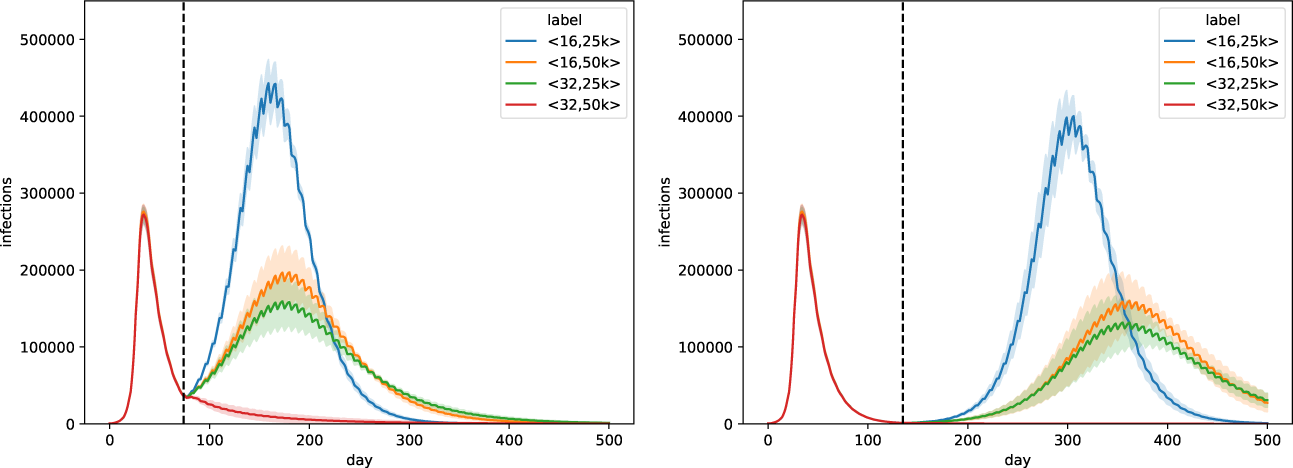
Trends for all combinations of parameters ⟨*k, T*_*d*_⟩, for FNR_PCR_ = 0.1. Universal testing starts at the first of May (left panel) and the first of July (right panel). We follow the pool isolation strategy, where we isolate all individuals that are part of an infected pool. The curves show a line that depicts the average over the trajectories of the result aggregations and a shaded area that depicts the standard deviation.

**Figure 19:**
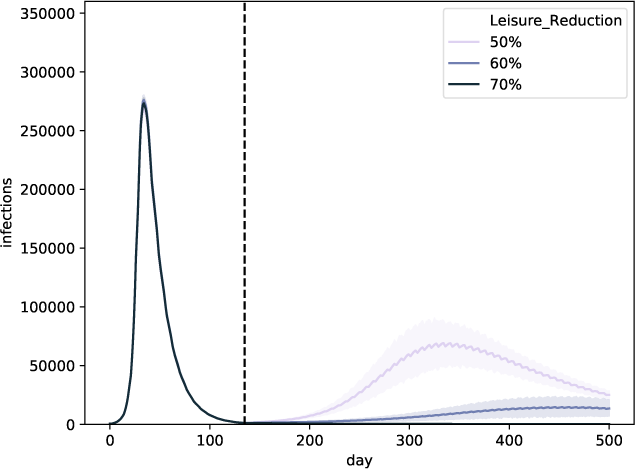
Trends for different leisure contact reductions, when performing weekly universal testing. We assume that universal testing starts on the first of July and that FNR_PCR_ = 0.1. We consider pool isolation and challenge the assumption of household isolation. The curves show a line that depicts the average over the trajectories of the result aggregations and a shaded area that depicts the standard deviation.

**Figure 20:**
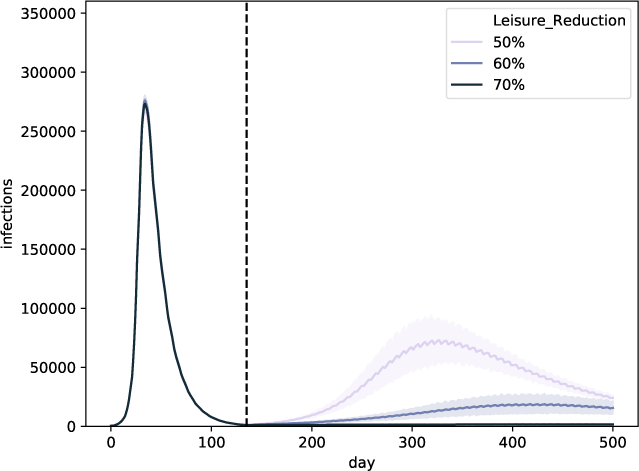
Trends for different leisure contact reductions, when performing weekly universal testing, and importing 10 cases per day. We assume that universal testing starts on the first of July and that FNR_PCR_ = 0.1. We consider pool isolation and challenge the assumption of household isolation. The curves show a line that depicts the average over the trajectories of the result aggregations and a shaded area that depicts the standard deviation.

**Figure 21:**
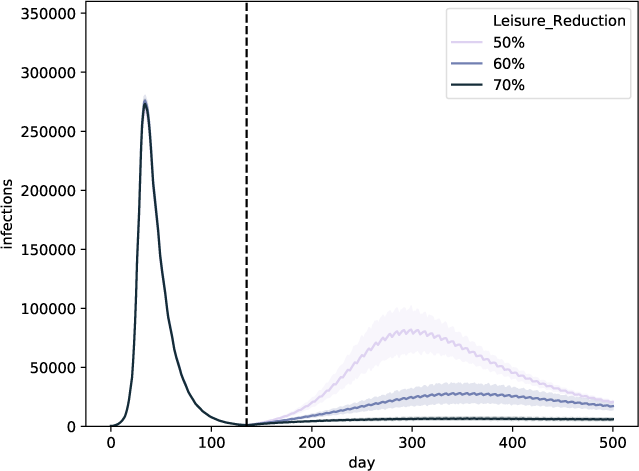
Trends for different leisure contact reductions, when performing weekly universal testing, and importing 50 cases per day. We assume that universal testing starts on the first of July and that FNR_PCR_ = 0.1. We consider pool isolation and challenge the assumption of household isolation. The curves show a line that depicts the average over the trajectories of the result aggregations and a shaded area that depicts the standard deviation.

## D PCR test reporting delay (*d*_*t*_) sensitivity analysis

**Figure 22:**
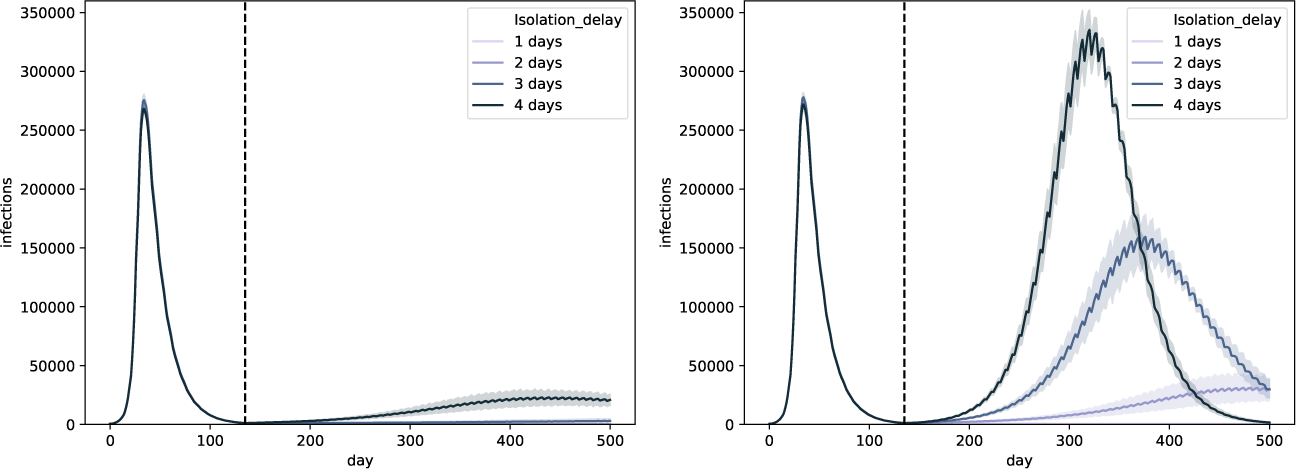
Sensitivity analysis for weekly universal testing (*k* = 32, *T*_*d*_ = 50k) with a contact reduction of 70%, considering *d*_*t*_ = {1, 2, 3, 4}. We assume that universal testing starts on the first of July and that FNR_PCR_ = 0.1. We consider both isolation strategies: pool isolation (left panel) and individual isolation (right panel). The curves show a line that depicts the average over the trajectories of the result aggregations and a shaded area that depicts the standard deviation.

**Figure 23:**
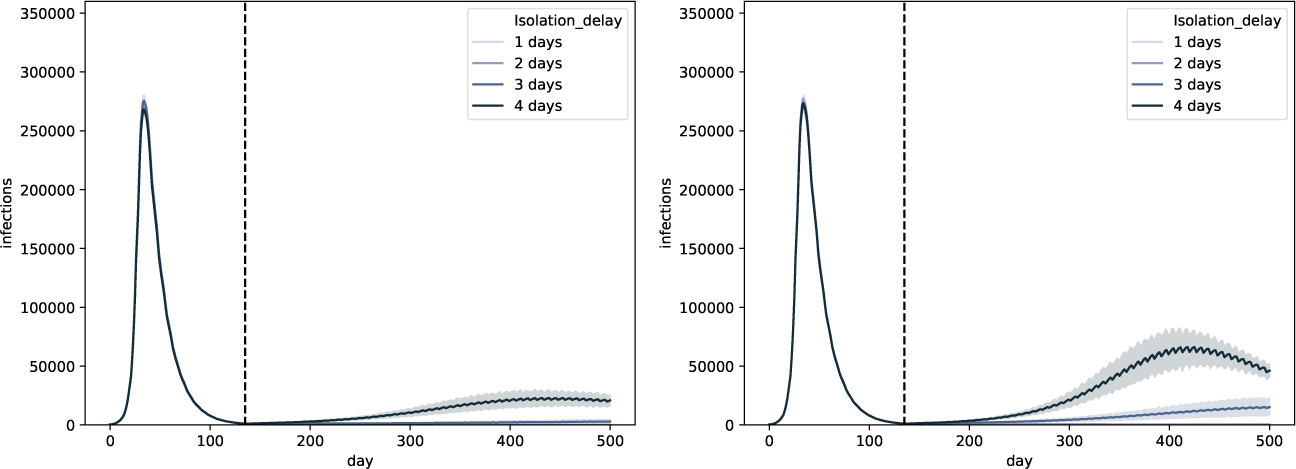
Sensitivity analysis for weekly universal testing (*k* = 32, *T*_*d*_ = 50k) with a contact reduction of 70%, considering *d*_*t*_ = {1, 2, 3, 4}. We assume that universal testing starts on the first of July and that FNR_PCR_ = 0.1. We consider both isolation strategies: pool isolation (left panel) and household isolation (right panel). The curves show a line that depicts the average over the trajectories of the result aggregations and a shaded area that depicts the standard deviation.

## E Weekly versus bi-weekly universal testing

**Figure 24:**
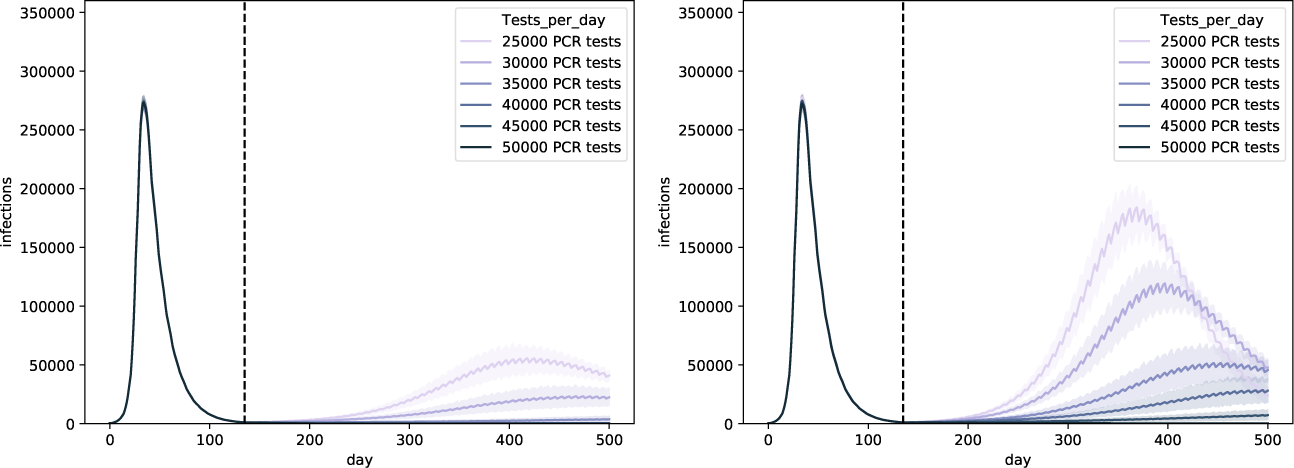
Weekly versus bi-weekly universal testing (i.e., *k* = 32,*T*_*d*_ ∈{25*k*, 30k, 35k, 40k, 45k, 50k}) with a contact reduction of 70%, considering *d*_*t*_ = {1, 2, 3, 4}. We assume that universal testing starts on the first of July and that FNR_PCR_ = 0.1. We consider both isolation strategies: pool isolation (left panel) and individual isolation (right panel). The curves show a line that depicts the average over the trajectories of the result aggregations and a shaded area that depicts the standard deviation.

## Footnotes

1 https://www.ecdc.europa.eu/en/geographical-distribution-2019-ncov-cases

2 We formulate a heuristic, as executing an optimal search algorithm is intractable. The reason is that this allocation setting is related to the bin-packing problem, which is NP hard [23].

3 https://ec.europa.eu/eurostat/web/nuts/background

4 We consider any infected individual to be an active case, which includes different symptomatic stages (pre-symptomatic, symptomatic, asymptomatic) and individuals that are not aware of their infection status.

5 k signifies kilo, i.e., 10^3^.

6 https://www.rapidtests.org

